# The single item physical activity (SIPA) measure: a major role for global surveillance and community program evaluation

**DOI:** 10.64898/2026.04.14.26350895

**Authors:** Adrian Bauman, Kat Owen, Sven Messing, Heather Macdonald, Lindsay Nettlefold, Giulia Coletta, Justin Richards, Corneel Vandelanotte, I-Hsuan Chen, Benny Cullen, Joe Van Buskirk, Anetta Van Itallie, Paul O’Halloran, Matthew Nicholson, Kiera Staley, Erica Randle, Heather McKay

**Affiliations:** School of Public Health, Faculty of Medicine and Health, Sydney University 2006 NSW Australia; Physical Activity for Health Research Centre, Health Research Institute, Department of Physical Education and Sport Sciences, University of Limerick, Limerick, Ireland; Active Aging Research Team, University of British Columbia, Vancouver, Canada and; Department of Family Practice Faculty of Medicine University of British Columbia, Vancouver, Canada; Te Hau Kori, Te Herenga Waka—Victoria University of Wellington, New Zealand/Aotearoa; Sport New Zealand / Ihi Aotearoa, Wellington, New Zealand/Aotearoa; Sport Ireland, Snugborough Road, Blanchardstown, Dublin, Ireland; School of Psychology and Public Health, La Trobe University, Melbourne, Victoria Australia; Monash University Indonesia, Banten, Indonesia; Department of Sport Science and Sport, Friedrich-Alexander-Universität Erlangen-Nürnberg, Erlangen, Germany; Physical Activity Research Group, Appleton Institute, Central Queensland University, Rockhampton, 4701, Queensland, Australia; S.P.O.R.T. Research Cluster, Central Queensland University, Rockhampton, 4701, Queensland, Australia; Centre for Sport and Social Impact, La Trobe University, Melbourne, Victoria Australia

**Keywords:** physical activity, surveillance, measurement, evaluation

## Abstract

**Introduction:** Physical activity is an important global health issue, yet surveillance and measurement methods vary considerably across countries and programs. This paper examines the use of the single-item physical activity (SIPA) measure through a comparative analysis of three representative national surveys and evaluates how well the SIPA reflects national estimates of ’meeting cutpoints for physical activity guidelines’. Finally, we assess the application of the SIPA in the evaluation of four large community-based physical activity programs.

**Methods:** We describe the SIPA distribution using national data from the Active New Zealand Survey, the Australian National Health Survey and the Irish Sport Monitor. We explore the relationship between the SIPA and existing physical activity measures of guideline attainment using Receiver Operating Characteristic (ROC) analysis, with Area Under the Curve (AUC) as the measure of fit. For the four community programs, we compare pre- to post-program changes, including effect sizes (Cohen’s δ).

**Results:** Across the three national datasets, mean SIPA values ranged from 2.68 to 3.31 days/week, with a bimodal distribution—people reported either zero or seven days of physical activity/week. A cutpoint at a threshold of ≥3 days/week provided optimal classification of guideline attainment (AUC >0.8 in all countries). Across the four large community programs, SIPA values increased by a typical 0.5–1.0 days/week, with 10–30% more participants meeting physical activity guidelines post-program.

**Conclusion:** SIPA offers a feasible, low-cost option for both population surveillance and community program evaluation. This is particularly relevant in low-income countries and settings, and complements growing interest in device-based measurement (e.g., accelerometry). Testing of SIPA in low- and middle-income countries is urgently needed. Despite the need for future research, SIPA holds immediate promise as a standardised physical activity measure for policymakers, researchers and community program evaluators.

**Key Messages:** There are many measures used for physical activity population surveillance and for large program evaluation, but the single item physical activity (SIPA) offers new potential for coordination and standardisation of measures used by asking adults about the number of days/week they were active. Limited research has described population use of the SIPA. This study describes population physical activity levels in national surveys in Australia, New Zealand and Ireland, and also describes the use of the SIPA in the evaluation of physical activity change in four large community physical activity programs. The SIPA provides a validated, comparable, and easy to administer measure for use in physical activity surveillance systems and for policymakers and practitioners evaluating community programs.

## Introduction

Physical activity is important for global health through its effects on reducing non-communicable disease risk, improving mental health and contributing to solutions to the climate change crisis (1–3). In general, physical activity research and policy approaches are under-represented in low resource settings. Hence the evidence-base is inequitable from a global perspective (4). One important public health task is population surveillance; for this, physical activity is most often assessed using self-report measures. These measures vary substantially between countries in the number of questions asked and the domains assessed. To illustrate, since 2002 the World Health Organization (WHO) recommended the use of the Global Physical Activity Questionnaire (GPAQ) as part of the non-communicable disease STEPwise surveillance program (5). Physical activity was assessed at least once in 122 mostly low-middle income countries (LMICs) through WHO STEPS surveys by the mid-2010s (5). However, since 2020, few countries reported GPAQ data through the STEPS surveys (based on an assessment of WHO reports).

Although physical activity measures are used for surveillance, in several countries, survey questions have changed over time as Changes were implemented as governments sought to capture new health thresholds or reflect new evidence, and to identify domain-specific estimates (e.g., leisure time exercise and sport; active travel). Importantly, changes to surveillance measures have disrupted the observed trends in the proportion of populations that meet physical activity guidelines (6, 7) .

Brief (1-2 questions) physical activity self-report measures are a pragmatic tool to assess physical activity in large samples/populations (8). They proved feasible for surveillance, but it was considered that they “*did not allow for measurement of whether an individual meets the World Health Organization (WHO) physical activity guidelines*” (8). Thus, the WHO and others (9, 10) are currently advocating for the use of device-based measures to standardise and improve population surveillance. However, only a few countries (all considered high-income) use device-based methods for national physical activity monitoring (11), and the fate of self-report surveys is not known. To exclusively use device-based methods more broadly in future may be limiting for a few reasons. Participation rates (selection effects) are low especially in low resource settings (12). International transition to device-based measures would be expensive, and serial data on physical activity levels acquired using existing self-report measures in representative samples would be lost.

Our specific single item physical activity (SIPA) question was developed in 2011, is a simple, feasible alternative (or complementary measure) to device-based measures for surveillance and for the evaluation of large-scale community programs (13) . SIPA has demonstrated reasonable repeatability (test-retest reliability coefficients of 0.7 to 0.8 (13, 14) and validity against accelerometers (correlations between 0.4-0.57), which was replicated in three language groups in Switzerland (15). It also demonstrated acceptable measurement properties when used to assess older adults in Canada (16) and has demonstrated responsiveness in interventions (17). Thus, SIPA has shown reliability and validity coefficients typical or better than more extensive self-report physical activity instruments in adults or older adults (18, 19).

Another advantage of short physical activity measures (including SIPA) is their potential to assess the effects of scaled-up and large community-based physical activity programs, comprising a broad range of participants across cultural or geographic settings (20). SIPA may be one solution that fills this sizeable measurement gap.

Therefore, the objectives of this study are twofold; (i) to assess the use of SIPA in population surveillance, and (ii) to illustrate how SIPA has been used in large-scale community-based physical activity program evaluation. To address our first objective, we describe the prevalence of physical activity (SIPA; days per week) among adults using data from three representative population national datasets—Active New Zealand (ANZ); Sport Ireland; National Health Survey Australia. We also address the ancillary question, ‘How well does the SIPA represent the proportion of adults meeting the current WHO aerobic physical activity guidelines, and where is the optimal cutpoint?’ (that best reflects the WHO guideline) (21) . To address our second objective, we provide descriptive data from four at-scale community-based programs in Canada, Ireland and two programs in Australia that used SIPA to assess physical activity.

## Methods

### Representative national physical activity datasets that used the SIPA

We accessed three population-representative datasets (Active New Zealand (ANZ) survey; Sport Ireland, National Health Survey Australia) that used the SIPA as a national surveillance measure. Our specific SIPA question for adult respondents was: “*In the past week, on how many days have you done a total of 30 minutes or more of physical activity which was enough to raise your breathing rate? (This may include sport, exercise and brisk walking or cycling for recreation or to get to and from places but should not include household or physical activity that is part of your job)”* (*13*) . SIPA data are expressed as days/week, reported from zero to seven. The units ‘days/week’ may be more easily understood and processed by participants across countries compared with the more granular ‘minutes or hours/day’ of physical activity.

The ANZ survey initially used an alternative form of SIPA to assess a continuous random sample of New Zealand adults (2017-2020) (21) . The survey was updated to provide internationally comparable and validated SIPA data, reported here (2021-2023). Sport Ireland conducts annual population surveys of sport and physical activity participation among adults using the Irish Sport Monitor; SIPA was used to assess physical activity in 2023 (22). The National Health Survey in Australia, conducted by the Australian Bureau of Statistics, uses established Active Australia surveillance questions (23) to report the prevalence of Australian adults who meet physical activity guidelines; SIPA was used to assess physical activity in 2014 and 2017. In Australia, the SIPA was estimated in the National Health Survey as ‘How many days/week were you active for at least 30 minutes per day?’ (see Supplementary Figure 1).

### Assessing the best cutpoint for SIPA to represent the proportion of adults meeting the current WHO aerobic physical activity guidelines

We compared SIPA outcomes against each countries’ established self-report surveillance measure to determine and monitor whether participants met the WHO aerobic physical activity guideline (i.e., 150 minutes or more of MVPA per week (24). Australian SIPA data were compared to established estimate of those meeting guidelines, derived from the six-question Active Australia physical activity measure (23). In New Zealand, the ANZ survey asked a single time-based question to identify people meeting guidelines—’How much physical activity (hours, minutes) did you participate in during the past week?’ (21). In Ireland, the Irish Sport Monitor summed time spent in all activities to assess the proportion of people who met the WHO physical activity guidelines (25) . From the Irish Sport Monitor there were three options for analysing data compared to SIPA: 1. meeting the ‘150 minutes physical activity guideline’ through totalling activities and sport, 2. adding walking for recreation, or 3. adding walking for recreation or transport (Supplementary Material Figure 2).

We conducted receiver operator characteristics (ROC) analyses to determine the relationship between SIPA and existing physical activity surveillance questions used to determine population groups meeting the WHO physical activity guideline (criterion measure). Specifically, from the ROC curves, we sought to assess the fit between the test measure (SIPA) and the established measures used in each country to assess whether physical activity guidelines were met (26). ROC curves plot sensitivity against 1-specificity and report the area under the curve (AUC) to assess overall significance of the association. AUC scores above 0.8 are considered very good to excellent (26). Youden’s index (sensitivity + specificity -1) is a measure of overall fit of the test measure against the established threshold criterion. The point at which the Youden’s index is maximal may be used to identify the optimal cutpoint in days per week that best reflects the criterion measure (27). We used population weights for all three national data sets. This type of analysis is used to identify thresholds on a test measure (e.g., SIPA), in relation to an existing criterion measure (e.g., national data for meeting physical activity guidelines) (26). We used SPSS version 29 (https://www.ibm.com/spss) and Medcalc (www.medcalc.org) to conduct our analyses.

### Large-scale community-based programs that evaluated physical activity using SIPA

Based on its simplicity and feasibility, several large-scale community physical activity and sport programs used SIPA to assess changes in physical activity. Choose to Move (British Columbia, Canada) is an evidence-based, 3-month health-promoting physical activity and social connectedness program (https://choosetomove.ca). This free program was effectively implemented across British Columbia in community settings in partnership with organisations that have broad reach to older adults (e.g., BC Recreation and Parks Association; YMCA). Choose to Move has been scaled up and engaged more than 9000 older adults (2016-March 2025) (28). Choose to Move used SIPA to assess older adults’ physical activity across different geographic regions; change data were pre-post participation in Choose to Move (baseline to 3-months).

Across Australia, the *10,000 Steps* program is a free, evidence-informed multi-strategy physical activity program. *10,000 Steps* motivates individuals to set goals and track their steps as motivation to increase daily physical activity (10000steps.org.au/). *10,000 Steps* is delivered via an interactive web and mobile apps (iOS and Android) using digital behaviour change techniques . Project officers based at Central Queensland University provide implementation support via e-mail and phone (see 10000steps.org.au/about-us/). SIPA was collected at baseline from all registrants in 10,000 Steps.

Sport Ireland recommends the use of SIPA as a standardised tool to evaluate diverse regional and community sports programs delivered across the country. Through Sport Ireland, we accessed data from each community sport program that used SIPA to assess change pre-post program participation. Irish community programs targeted many different sports and recreation activities in diverse populations.

The Centre for Sport and Social Impact (CSSI, La Trobe University, Melbourne) used SIPA to assess physical activity in community sport programs in Victoria, Australia (2015-2022) using registration and end of program survey time points. Thirty-nine state and regional sport organisations were funded by the Victorian Health Promotion Foundation (VicHealth) to engage adults not meeting physical activity guidelines in new social sport programs. They targeted rural, disadvantaged and diverse communities, engaging more than 39,000 adults.

For all four community programs, we conducted descriptive analyses to illustrate the distribution of responses and changes in physical activity among individuals enrolled in community programs. We assessed change from baseline to the first follow up time point (physical activity change scores; days/week ± 95% confidence intervals (CI)) and Cohens δ effect sizes.

## Results

### Representative national physical activity datasets that used the SIPA

Population demographics were similar across national datasets: 51% (NZ), 51.2% (Ireland), and 50.8% (Australia) were female, and 44.2%, 42.4%, and 42.1%, respectively, were aged ≥50 years. We summarize SIPA responses for each national population survey (Figure 1). There were bimodal distributions for Australia and Ireland; survey participants reported 0 days active or 7 days active more often compared with other responses. Physical activity (mean days/week) were 3.31 (NZ), 3.07 (Ireland) and 2.68 (Australia) (Table 1).

**Figure 1.**
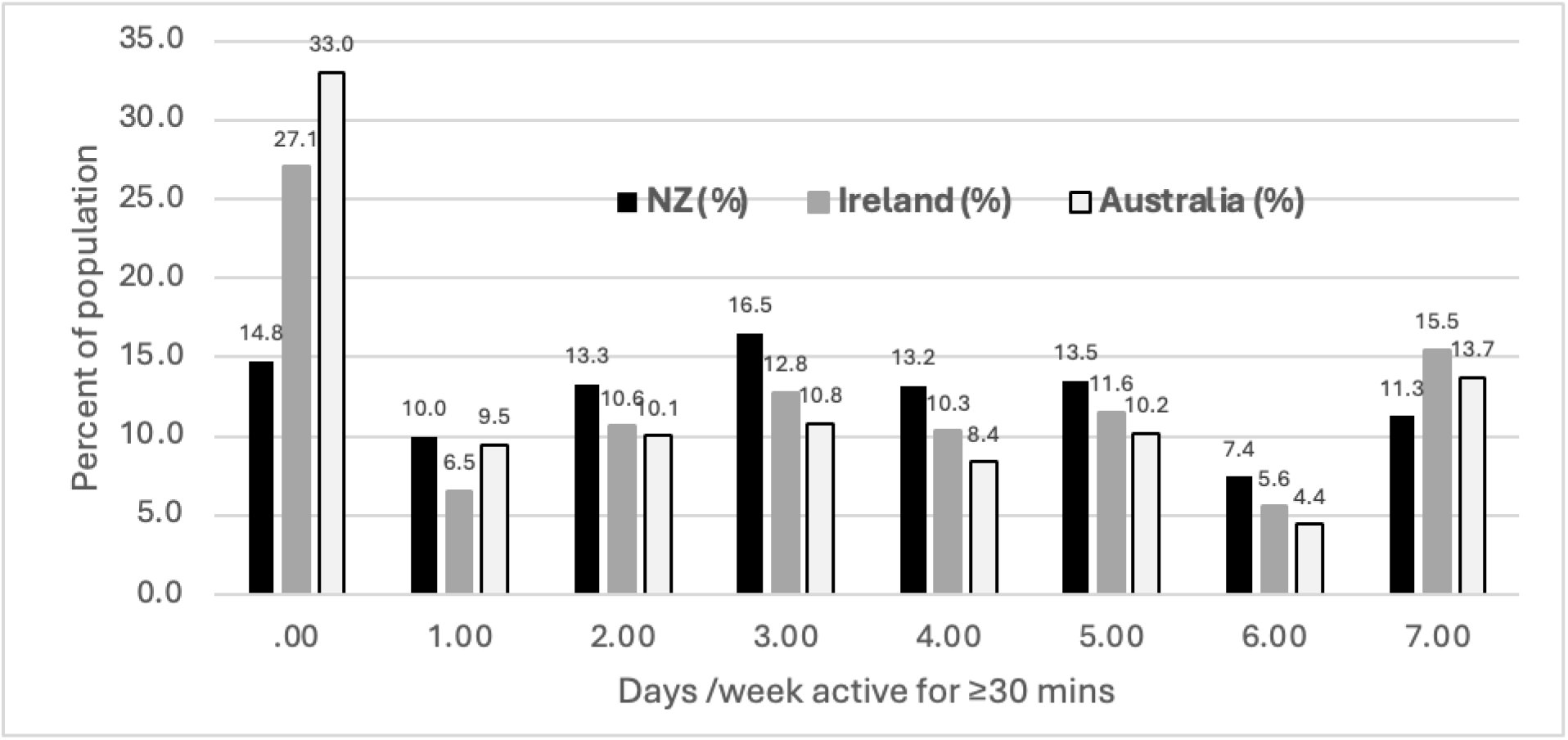
Proportion of adults reporting 0 to 7 days/week of 30 minutes or more physical activity as measured with the single-item physical activity measure, in national surveys (New Zealand, Ireland, Australia).

**Table 1.** Using the single item physical activity (SIPA) question to assess the number of days/week that best represents meeting PA guidelines (≥150 min/week) compared with established self-report surveillance measures for meeting the physical activity guideline (150 mins/week)

| Country population dataset | Single item physical activity descriptive data, mean days/week (sd) | Established physical activity measure for comparison with SIPA<sup>ϕ</sup> | Area under the ROC curve (95%CI) * | Youden's index (95%CI) <sup>θ</sup> | Best fit<sup>¶</sup> |
| --- | --- | --- | --- | --- | --- |
| Active New Zealand / Aotearoa 2021-23<br>N=53961 | 3.31 (2.21) | Total reported time/week in all PA ($\geq 150$ mins) | 0.81 (0.81-0.82) | 0.54 (0.52-0.55) | $\geq 3$ days+/week |
| Irish Sport Monitor 2023<br>N=8423 | 3.07 (2.51) | Sum of time in all activities + walking for recreation or transport ($\geq 150$ mins) | 0.84 (0.83-0.85) | 0.55 (0.53-0.57) | $\geq 3$ days+/week |
| Australian NHS 2014 and 2017 <sup>Ω</sup><br>N=30930 | 2.68 (2.58) | Active Australia ($\geq 150$ mins) Mod PA + Vig PA + walking | 0.93 (0.92-0.97) | 0.71 (0.67-0.73) | $\geq 3$ days+/week |

**LEGEND** <sup>ϕ</sup> compared to established self-report question for assessing meeting PA guidelines
\* Area under the curve (c) derived from ROC curve plot of Sensitivity against (1-Specificity)
<sup>θ</sup> Youden's index = Se + Sp – 1 (measure of best fit)
<sup>¶</sup> point of best fit, least misclassification as it maximises Sensitivity and Specificity of SIPA compared to the established PA measure
<sup>Ω</sup> in the Australian Bureau of Statistics National Health Survey asked as two questions (see Supplementary material 1) but the question reported here includes days/week of 30 minutes/day, comparable with ANZ and Irish SIPA data

In Australia and New Zealand, 44.1% and 51.7% of adults, respectively, met physical activity guidelines using existing surveillance measures. In Ireland, 31.4% of adults met physical activity guidelines through sport alone; the proportion increased to 61.3% if recreational walking was added, and to 69.4% if walking for both recreation and transport were included.

We summarise the results from the ROC analysis for each country (Table 1); curves compare the SIPA to existing surveillance measures. All area under the curve (AUC) statistics were >0.8. Physical activity data from Australia showed AUCs >0.9. Youden’s index that maximised sensitivity and specificity were very good to excellent. We provide ROC curves in Supplementary Figure 5. The optimal cutpoint for classification as ‘meeting guidelines’ in each country was at least 3 days/week using the SIPA (Table 1). Additional ROC analyses by age group, gender and socio-economic measures are provided in Supplementary Table 1 and show very similar AUC values across demographic subgroups in all three countries.

### Large-scale community-based programs that evaluated physical activity using SIPA

Table 2 shows SIPA data from four community programs. As providing baseline data was often mandatory at sports or physical activity program enrolment, baseline sample sizes were often much larger than those at (voluntary) follow up. The distribution of SIPA responses in the community program evaluation samples differed from the three national population datasets. For all four community programs,, a large proportion of the population reported 2-3 days/week of physical activity at baseline (Table 2). Choose to Move in Canada specifically recruited a sample of low active older adult participants. Physical activity at first follow-up increased 0.95 days (*10,000* steps), 0.54 days (Ireland), 1.2 days (Choose to Move) and 0.15 days (Sport Victoria). The first three of these reflect medium effect sizes using Cohen’s δ; the largest effect size was in the more structured Canadian Choose to Move intervention (Table 2). The proportion of the samples meeting physical activity guidelines based on the ROC-defined 3+days/week threshold increased by 10.4% (95% CI 8.9-11.9; 10,000 Steps), 11.8% (CI 10.5-13.1; Ireland), and 28% (CI 25.8-30.1; Choose to Move). Changes were smaller for the Victorian sports programs (2.9%; CI -0.5 – 5.8%). When we applied a 5+ days/week threshold, increases in physical activity were 20.4%, 8.7%, 17% and 1.1% for *10,000* steps, Irish community sport programs, Choose to Move, and Victorian sports programs, respectively.

**Table 2.** Single item physical activity data (SIPA days/week) from community sport and physical activity programs.

| **Program and type of data** | **10,000 Steps, Australia** | **Sport Ireland community programs** | **Choose to Move, British Columbia, Canada** | **Sport Victoria community programs, Australia** |
| --- | --- | --- | --- | --- |
| Type of program | Community-wide digital physical activity program with multi-strategy implementation resources; national reach | Variety of community sport and activity programs for all age groups in all regions | Supported physical activity and social connectedness program for older adults; scaled up across British Columbia, Canada | Thirty-nine organisations developed, tested and/or supported delivery of social sport programs for adults (18+ years) in Victoria, Australia |
| Single item PA baseline days/week; mean (sd); n | 3.53 (1.11); n=135465 | 3.47 (1.97); n=14170 | 2.3 (2.0); n=4864 | 3.31 (1.9); n=21,413 |
| Distribution of baseline responses by days/week active for $\geq 30$ mins (%) | | | | |
| 0 days % | 7.1 | 2.2 | 25.0 | 8.1 |
| 1 day | 7.7 | 5.7 | 14.3 | 13.2 |
| 2 days | 13.3 | 12.6 | 18.4 | 18.4 |
| 3 days | 20.1 | 22.8 | 17.4 | 22.0 |
| 4 days | 15.0 | 17.7 | 9.3 | 14.7 |
| 5 days | 16.5 | 17.7 | 6.8 | 11.5 |
| 6 days | 6.1 | 8.9 | 3.4 | 5.1 |
| 7 days | 14.2 | 12.5 | 5.4 | 7.0 |
|  | *N=6238* | *N=8323* | *N=4864* | *N=21413* |
| Pre/post program means (sd) and initial change score (d) (days/week) to first follow up | Pre: 3.73 (2.05)<br>Post: 4.68 (2.15)<br>Change ($\Delta$): 0.95 (2.22)<br>Cohen's $\delta$: 0.45 (0.42-0.49)<br>Matched sample n=6237 | Pre: 3.46 (1.91)<br>Post: 3.99 (1.80)<br>Change ($\Delta$): 0.54 (1.60)<br>Cohen's $\delta$: 0.28 (0.26-0.29)<br>Matched sample n=7028 | Pre: 2.3 (2.0)<br>Post: 3.5 (2.0)<br>Change ($\Delta$): 1.2 (2.1)<br>Cohen's $\delta$: 0.60 (0.55-0.65)<br>Matched sample n=3769 | Pre 3.12 (1.9)<br>Post 3.27 (1.8)<br>Change 0.15 (1.64)<br>Cohens $\delta$ 0.08 (0.03-0.26)<br>Matched sample n=2081 |
| Pre/post % meeting $\geq 3$ days/week active (matched samples) | Pre: 71.9%<br>Post: 82.3% | Pre: 67.7%<br>Post: 79.5% | Pre: 42%<br>Post: 70% | Pre (n=2081) 60.4%<br>Post: 63.3% |
| Pre/post % meeting $\geq 5$ days/week active (matched samples) | Pre: 36.8%<br>Post: 57.2% | Pre: 30.4%<br>Post: 39.1% | Pre: 15%<br>Post: 32% | Pre (n=2081) 23.6%<br>Post: 24.7% |

Only a small proportion of participants who enrolled in the *10,000* Steps, Irish and Victorian programs had a follow up assessment. It is not known what that number is for Ireland, as myriad community programs potentially used the SIPA. For Choose to Move, a structured intervention, most participants consented to be evaluated at baseline and 3-month follow-up. For *10,000* steps, we assessed differences in physical activity (SIPA; days/week) between baseline registrants (n=135465) and the sample at follow up (n=6238). Distributions in physical activity were similar between groups (Supplementary Figure 4).

## Discussion

We demonstrate that a simple, single item physical activity measure (SIPA) is a feasible option to assess physical activity for population surveillance and to evaluate community-based physical activity programs at low cost among adults and older adults. For surveillance, we used representative population-based surveys from three different countries and demonstrated similar distributions of physical activity (days/week) across surveys. Across all three countries, the SIPA could, with good sensitivity and specificity, estimate the proportion of adults who met the aerobic physical activity guidelines. The optimal cutpoint was 3+ days/week.

The WHO 150 minutes/week physical activity guideline is still characterised in some countries as ‘150 minutes, 5 times per week’ or ‘5x30 minutes of activity/week’ (29, 30) . Research can sometimes influence policy change, as the findings prompted changes in how these data from Sport New Zealand were interpreted; researchers using these SIPA results moved from reporting 5+ days/week as meeting PA recommendations to adopting the 3+ days/week threshold (21). This is an important (more achievable) distinction, given the challenges confronting adults and older adults to meet recommended guidelines for physical activity. Our analyses in all three countries support the SIPA-derived cutpoint of 3+ days/week of physical activity to meet guidelines. Policymakers and physical activity surveillance systems might consider adopting this evidence-supported, more achievable goal.

In physical activity research, ROC analyses have been used to examine differences between device-based measures or to define physical activity intensity cutpoints (31, 32). However, ROC analyses used less often for population surveillance, despite their utility. More than a decade ago, Zwolinsky and colleagues reported poor correlations between SIPA and IPAQ (33). They suggested that ROC analyses were most useful to classify inactive individuals. A recent study of 55,000 European adults adopted the ROC approach to evaluate whether a single-item physical activity question was an adequate substitute for much longer physical activity questionnaires . They used the well-established International Physical Activity Questionnaire (IPAQ) as the test measurement to predict a categorical version of SIPA as the outcome. They showed good to excellent AUC values (0.75-0.95). However, our study may be more relevant to global surveillance needs, as we examined SIPA as the new test measure and compared it to established longer self-report measures currently used to assess physical activity guidelines.

Four large community-based programs demonstrated that SIPA was a simple, feasible tool that could be used to measure change in physical activity in sport and other health promoting programs. SIPA has demonstrated good measurement and validity properties, comparable to the usual and more extensive physical activity surveys of adults (13, 14) and older adults (16) . This is especially important in large scale-up studies and low resource settings where costs for device-based measures or collection of multi-question physical activity measures are prohibitive. Notably, the SIPA was sensitive enough to capture change in community interventions (17) . Thus, effect sizes can be determined and stratified by demographic sub-groups and by different types of community programs. These data can then be used to develop evidence-based targets. The policy implications are that government investments in community programs, decision making and messaging to the public would be grounded in evidence.

The distribution of SIPA in community samples differed from general population data (Supplementary Figure 3). This is expected given that participants in community programs often engage in some level of physical activity. They often enrol in programs with the goal of increasing their physical activity levels. This is not the case with population data which is more bimodal--a greater proportion of people either report doing no physical activity or report being active every day.

Among the four community programs, the Choose to Move program in Canada was a community-wide intervention, co-designed based on evidence (behaviour change principles) to enhance physical activity. A central support team provided ongoing support to delivery partner organisations and encouraged participants to engage in follow-up measurements at the end of the program. By contrast, the other community programs were less formalised and diverse. Data were useful but were acquired on only a small proportion of participants enrolled at baseline.

### Limitations and strengths

We acknowledge that our study had a number of limitations. Given that SIPA-based measures were reported in few large scale programs to date, our data come from high-income countries. Another example was reported by Sport England, where SIPA was used to screen for low active individuals to prioritise access to community sport programs (34). This review indicated that most programs used more complex measures (e.g., IPAQ) to evaluate changes in physical activity over time. However, stakeholders reported that the longer IPAQ measure was often not a practical or feasible evaluation tool (34). The perspective of stakeholder groups is important—they serve as implementation partners and/or often fund the evaluation of community based physical activity programs. Another limitation is that the SIPA lacks context and domain-specific estimates, as these estimates can only be gleaned from more comprehensive physical activity measures. This problem is also common to device-based surveillance.

Although the Irish sport programs (3 months post) and Choose to Move from Canada (3 or 6, and 12 months post) collected longer term follow-up, relatively few community-based participants in our studies provided SIPA post-program data. However, it was not our intent to test physical activity effects over the longer term or to demonstrate representativeness of community program evaluations. Our purpose was to ascertain whether the SIPA was a feasible option to assess the immediate impact of community physical activity programs. Loss to follow-up is a concern for many programs delivered to large populations. Strategies that decrease attrition should be considered; improved follow-up data would increase the generalisability of our findings.

Field testing of the SIPA, especially in low-resource communities, is an important next step. If device-based surveillance becomes the sole method used to assess physical activity at scale, countries may cease collecting their usual self-report surveillance measures (such as IPAQ or GPAQ). It is therefore important that we determine SIPA measurement properties and responsiveness in population surveillance and for community program evaluation in low-income populations, in low-middle income countries and in languages other than English. To date, only one validation study was conducted in German, Italian and French (15). We conducted equity-focused analyses (Supplementary Table 1) and showed that AUC values were very similar by age group, gender and socioeconomic status. Thus, the utility of the SIPA did not differ by population subgroups in the three high-income countries in our study. The national studies used different surveillance questions to estimate meeting physical activity guidelines. This may be a strength, as the SIPA performed similarly against all of them—using SIPA a physical activity cutpoint of 3+ days was recommended for adults to meet guidelines.

Overall, the strength of the SIPA is that it is a low cost, low burden, sustainable tool that can be utilised in population surveillance systems with measurement properties comparable to other self-report measures. The SIPA could be used as a standardised measure to replace or complement data acquired from device-based surveillance tools. The SIPA is useful for policymakers to compare the effects of engaging in diverse settings for health promotion and sports programs. Community-based program participants could also be compared with national norms from population-wide representative surveys. The SIPA is a flexible tool that can be adapted to use in other settings such as clinical populations, in primary care (8) and for populations with special needs (35, 36) . The SIPA is practical and could be easily embedded and sustained in national health surveys, irrespective of other changes to physical activity surveillance. Finally, the SIPA is an efficient option that demands much less survey time than more detailed questionnaires or device-based physical activity measures.

## Conclusion

Physical activity researchers, funding agencies, and community partners have for many years, sought a simple, affordable, robust and standardised way to assess physical activity. Indeed, a way to standardise measurement of physical activity has been the ‘holy grail’ of measurement for many decades (7). The SIPA may provide a long-term, low-cost solution, that could be used in large scale-up studies, low resource contexts and for sustained and comparable population surveillance. The SIPA could also be used to compare effects across programs and groups. It also seems pertinent (and feasible) to use the SIPA in community programs, and to embed it in population surveys alongside existing measures. Concurrent use of existing physical activity measures and the SIPA [and device-based tools] would enable local calibration and provide an opportunity to compare trends across populations and different tools. We contend that the initial intent of the SIPA (13) - to evaluate ‘real world’ low resource community physical activity programs - is laudable as it provides an equitable option to assess physical activity programs across many different contexts.

## Author’s contributions

*AB* conceptualisation [C], design [D], analysis [A], coordination and management[CM], writing draft [WD], writing and editing final version [WF], submission [S]

*Authors KO,SM,JR,IHC,BC,KS,ER,MN,HM : C,D,WD,WF*

*Authors POH,CV,AVI, JVB, HM, LN,GC: C,D,A WD,WF*

## Data sharing statement

technical details of data available on request; each data set requires its own permission from the data custodian

## Data Availability

All data are available subject to the rules for each custodian in the seven data sets used [for example, approaches to the Australian government, to Sport Ireland or Sport New Zealand), or to the custodians of the four community interventions [all are authors on this paper and could be approached for data access]

## Acknowledgements

We thank the data custodians for access to population data (i.e., Active New Zealand Survey, the Australian National Health Surveys and the Irish Sport Monitor). We are also grateful to colleagues who provided data from community-based interventions: 10000 STEPS, Choose to Move, Sport Ireland and the Victoria Community Sports programs and to the various partners who supported these studies, and community members who provided these data.

---

**Supplementary Figure 1.**
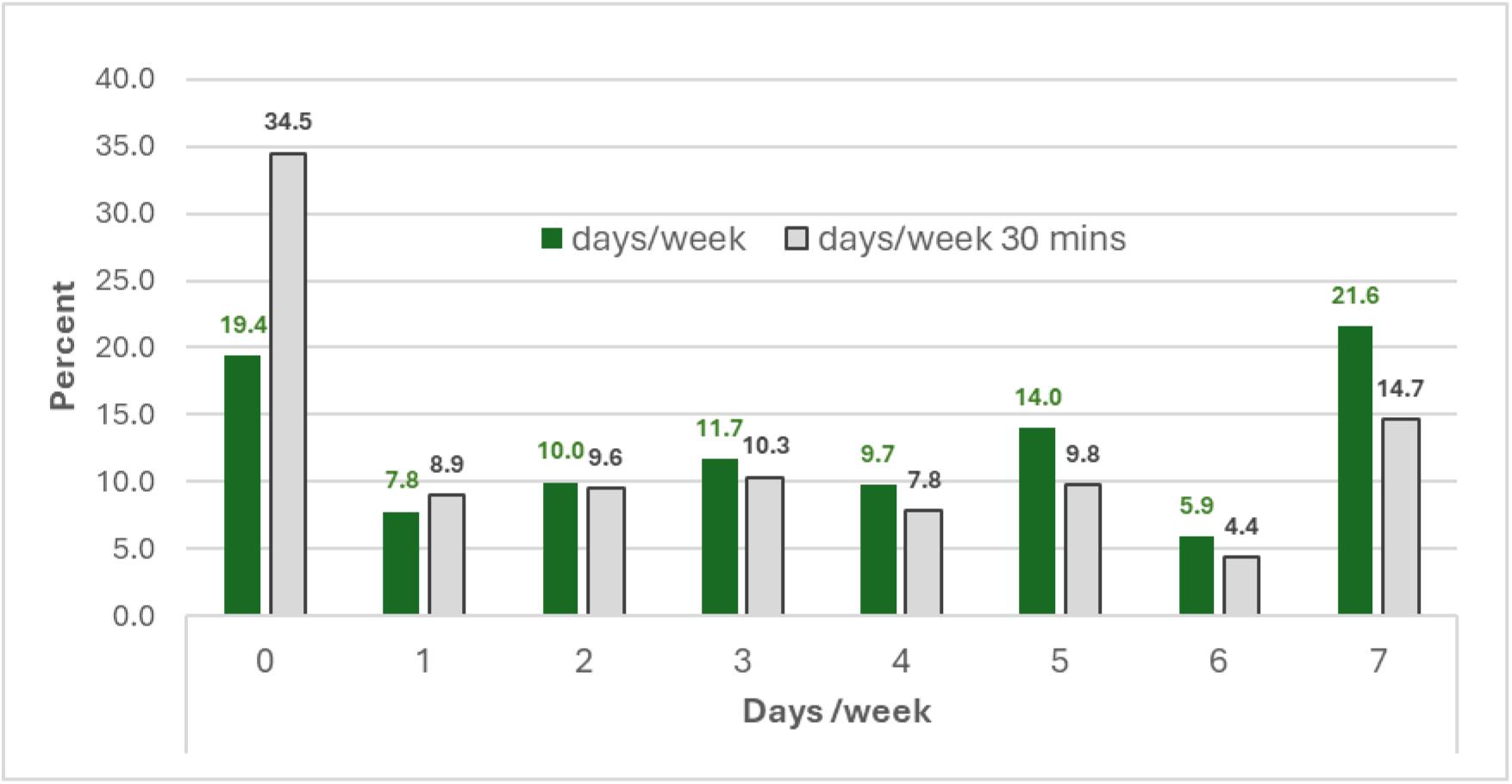
Comparison of Australian National Health Survey asked about physical activity (days/week) and the second question asked about physical activity (days/week that were of 30+mins’ (%): *Note that in the Australian National Health Survey, two separate single item questions were asked, the first asked only about physical activity as days/week, and the second asked about physical activity as days/week for at least 30 mins on those days; the latter is the exact SIPA question comparable with Irish and New Zealand SIPA questions.

**Supplementary Figure 2.**
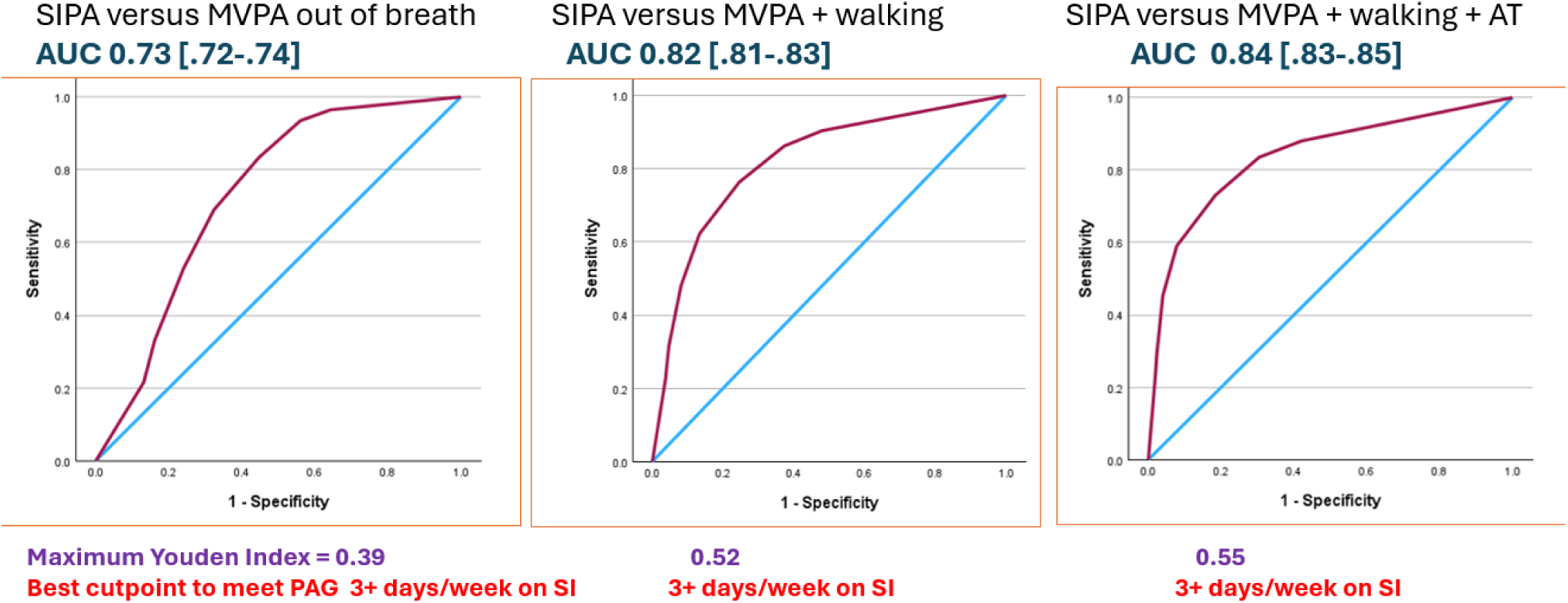
Irish data: SIPA compared to three established measures of physical activity among Irish adults: <u>Note:</u>. the SIPA includes all sport, exercise and recreational activity, but also mentions active travel (walking /cycling to from places); hence it is not surprising that using only the prevalence of sport participation (left hand panel) SIPA shows moderate utility, but adding recreational walking (middle panel) and adding active travel walking (right hand panel) improves the fit of the SIPA in identifying those meeting the 150 minute/week PA guideline.

**Supplementary Figure 3.**
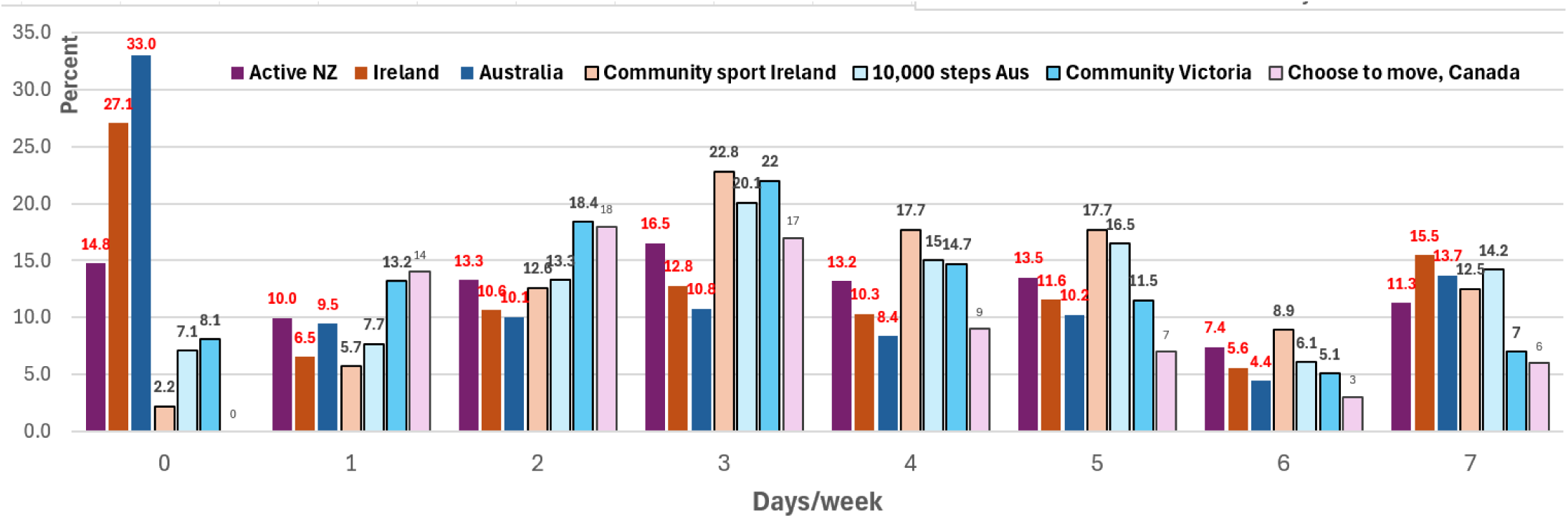
Comparison of national population data (dark bars) and community-based program baseline physical activity data (light colour bars) against the distribution of the single item physical activity (SIPA) measure (%)

**Supplementary Figure 4.**
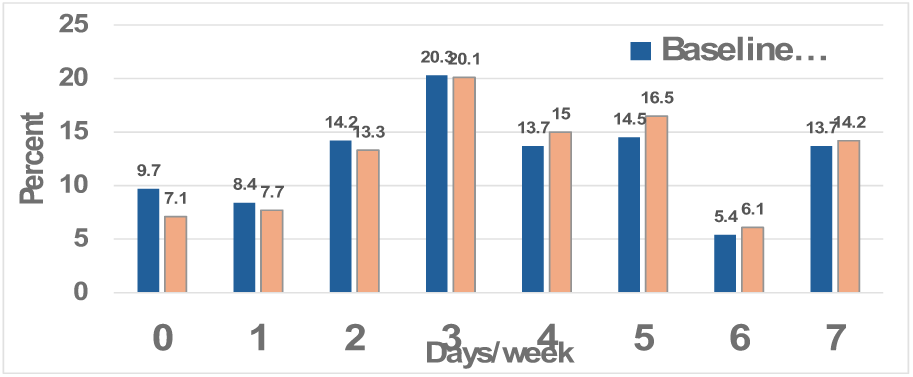
Community wide program, *10,000* Steps Australia – compares distribution of SIPA data for the large baseline mandatory registration sample (n=135465) to the smaller follow-up sample (n=6238) (%, days/week)

**Supplementary Figure 5.**
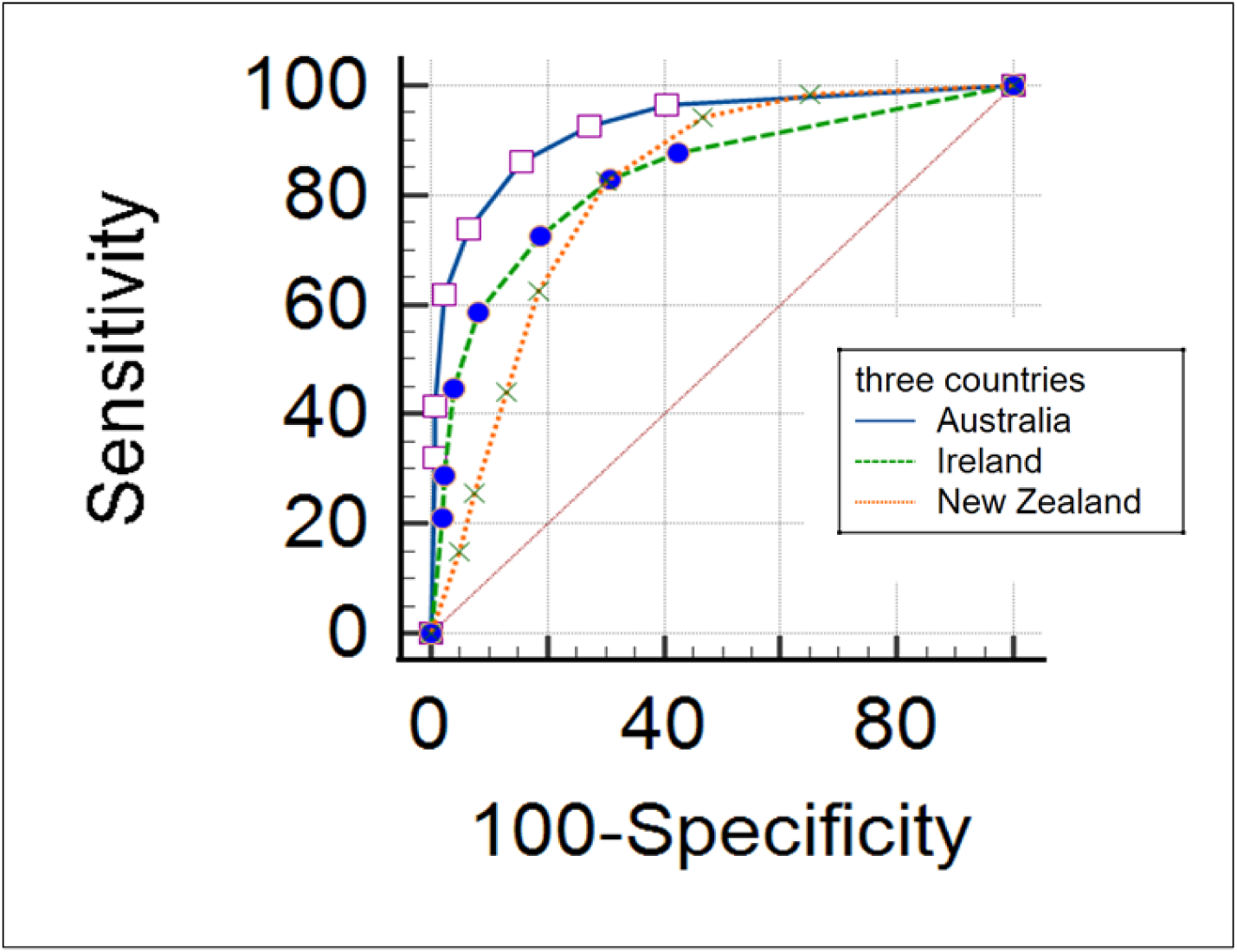
The ROC curves for physical activity (days/week) from the SIPA against meeting PA guidelines. Data are national population data from three countries*: Note: for all three countries combined, the AUC = 0.86 (0.85-0.87), Youden’s index= 0.58

**Supplementary Table 1.**
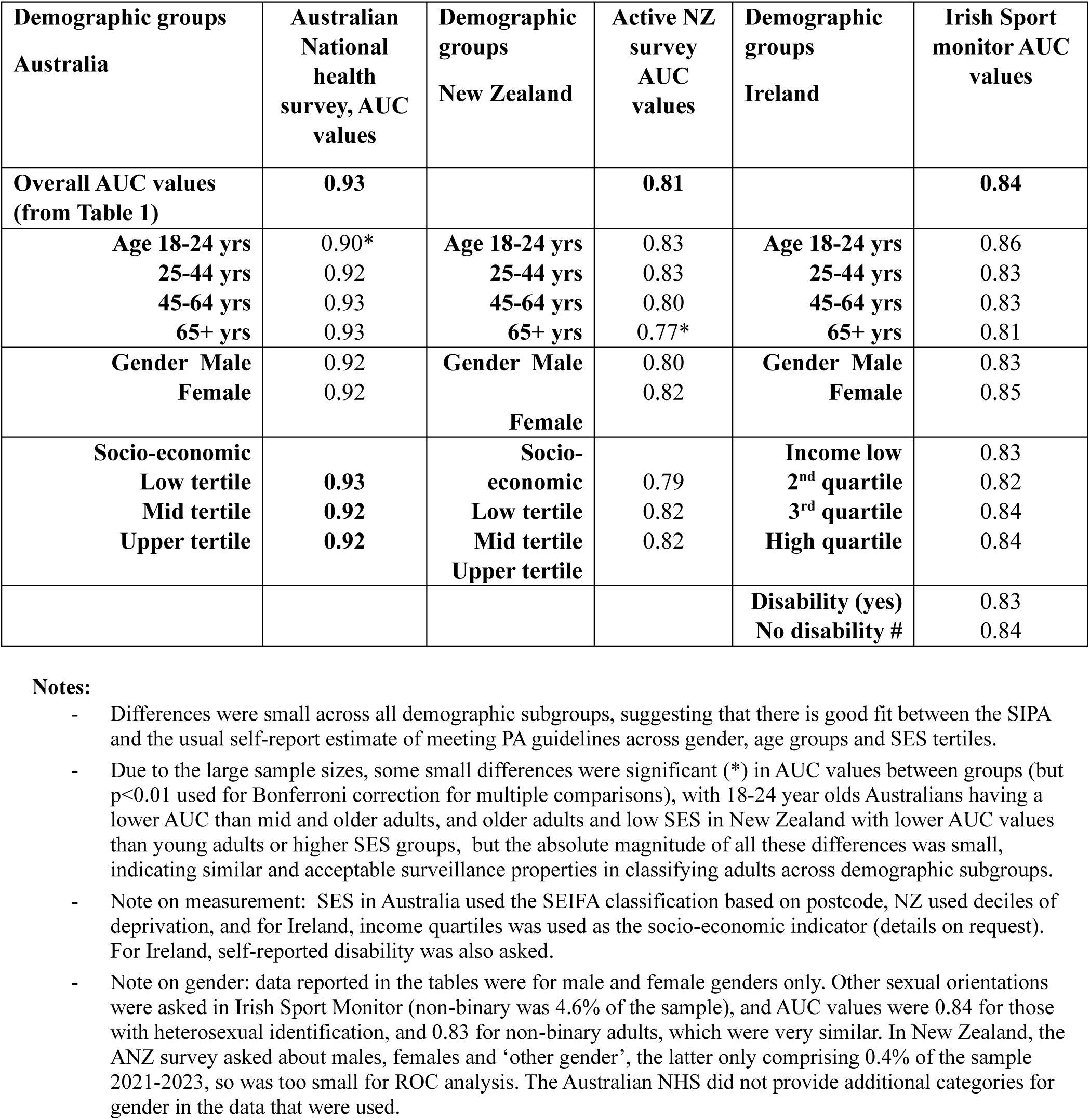
Area under the curve values from the ROC analyses of population data sets from Australia, Ireland and New Zealand. Data are stratified by age, gender and socio-economic status.

## References

1. Ramírez Varela A, Bauman A, Woods CB, Shawar YR, Hallal PC, Salvo D, et al. Low global physical activity despite two decades of policy progress. Nature Health. 2026;1(3):338–54.

2. Salvo D, Crochemore-Silva I, Wendt A, Tarp J, Shiroma EJ, Simpson RJ, et al. Physical activity for public health in the 21st century. Nature Medicine. 2026:1–11.

3. Hinckson E, Reis R, Romanello M, Ding D, Adelekan I, Favarão Leão AL, et al. Benefit of physical activity initiatives for climate change mitigation and adaptation. Nature Health. 2026;1(3):300–15.

4. Ramírez Varela A, Bauman A, Woods CB, Shawar YR, Hallal PC, Salvo D, et al. Physical activity remains under-prioritized in political agendas. Nature Health. 2026;1(3):278–9.

5. Riley L, Guthold R, Cowan M, Savin S, Bhatti L, Armstrong T, et al. The World Health Organization STEPwise approach to noncommunicable disease risk-factor surveillance: methods, challenges, and opportunities. American journal of public health. 2016;106(1):74–8.

6. Milton K, Bauman A. A critical analysis of the cycles of physical activity policy in England. International Journal of Behavioral Nutrition and Physical Activity. 2015;12(1).

7. Bauman A. Trends in exercise prevalence in Australia. Community health studies. 1987;11(3):190–6.

8. Tcymbal A, Messing S, Mait R, Perez RG, Akter T, Rakovac I, et al. Validity, reliability, and readability of single-item and short physical activity questionnaires for use in surveillance: A systematic review. PLoS ONE. 2024;19(3 March).

9. WHO. Physical activity measurement and surveillance in adults: report of a scoping and planning meeting, 27-28 November 2023. World Health Organization, Geneva 2024. Report No.: ISBN 978-92-4-009555-7

10. Troiano RP, Stamatakis E, Bull FC. How can global physical activity surveillance adapt to evolving physical activity guidelines? Needs, challenges and future directions. British Journal of Sports Medicine. 2020;54(24):1468–73.

11. de Wolf I, Elevelt A, van Nassau F, Toepoel V, de Hollander E, Kompier ME, et al. Comparing national device-based physical activity surveillance systems: a systematic review. International Journal of Behavioral Nutrition and Physical Activity. 2024;21(1).

12. Christiansen LB, Koch S, Bauman A, Toftager M, Bjørk Petersen C, Schipperijn J. Device-based physical activity measures for population surveillance—issues of selection bias and reactivity. Frontiers in sports and active living. 2023;5:1236870.

13. Milton K, Bull FC, Bauman A. Reliability and validity testing of a single-item physical activity measure. British Journal of Sports Medicine. 2011;45(3):203–8.

14. Milton K, Clemes S, Bull F. Can a single question provide an accurate measure of physical activity? British Journal of Sports Medicine. 2013;47(1):44–8.

15. Wanner M, Probst-Hensch N, Kriemler S, Meier F, Bauman A, Martin BW. What physical activity surveillance needs: validity of a single-item questionnaire. British Journal of Sports Medicine. 2014;48(21):1570–6.

16. Macdonald HM, Nettlefold L, Bauman A, Sims-Gould J, McKay HA. Pragmatic Evaluation of Older Adults’ Physical Activity in Scale-Up Studies: Is the Single-Item Measure a Reasonable Option? Journal of Aging and Physical Activity. 2022;30(1):25–32.

17. O’Halloran P, Sullivan C, Staley K, Nicholson M, Randle E, Bauman A, et al. Measuring change in adolescent physical activity: Responsiveness of a single item. Plos One. 2022;17(6).

18. van Poppel MN, Chinapaw MJ, Mokkink LB, van Mechelen W, Terwee CB. Physical activity questionnaires for adults: a systematic review of measurement properties. Sports Medicine. 2010;40(7):565–600.

19. Sattler MC, Jaunig J, Tösch C, Watson ED, Mokkink LB, Dietz P, et al. Current evidence of measurement properties of physical activity questionnaires for older adults: an updated systematic review. Sports Medicine. 2020;50(7):1271–315.

20. Mudiyanselage D BC, Chappel SE I, S, Fisher G, Crozier, AJ, Vandelanotte C Scoping review to assess the reach, effectiveness, and impact of government-funded, population-based physical activity initiatives in Australian adults. Frontiers in Sports and Active Living. 2025;7:1633086.

21. Bauman AE, Richards JA. Understanding of the Single-Item Physical Activity Question for Population Surveillance. Journal of Physical Activity and Health. 2022;19(10):681–6.

22. Lacey E, Cullen B. 217 The Irish Sports Monitor 2011-2023: Data Driven Insights-The Story So Far. European Journal of Public Health. 2024;34(Supplement_2):ckae114. 025.

23. AIHW. The Active Australia Survey: a guide and manual for implementation, analysis and reporting. https://www.aihw.gov.au/reports/physical-activity/active-australia-survey/summary. Canberra; 2003.

24. Ding D, Mutrie N, Bauman A, Pratt M, Hallal PR, Powell KE. Physical activity guidelines 2020: comprehensive and inclusive recommendations to activate populations. The Lancet. 2020;396(10265):1780–2.

25. Sport Ireland. Irish Sport Monitor 2022. Sport Ireland, Dublin, Ireland 2022.

26. Nahm FS. Receiver operating characteristic curve: overview and practical use for clinicians. Korean journal of anesthesiology. 2022;75(1):25–36.

27. Stanaway FF, Gnjidic D, Blyth FM, Le Couteur DG, Naganathan V, Waite L, et al. How fast does the Grim Reaper walk? Receiver operating characteristics curve analysis in healthy men aged 70 and over. BMJ (Online). 2011;343(7837).

28. McKay HA, Nettlefold L, Sims-Gould J, Macdonald HM, Khan KM, Bauman A. Status Quo or Drop-Off: Do Older Adults Maintain Benefits From Choose to Move-A Scaled-Up Physical Activity Program-12 Months After Withdrawing the Intervention? Journal of Physical Activity & Health. 2021;18(10):1236–44.

29. Marques A, Sarmento H, Martins J, Nunes LS. Prevalence of physical activity in European adults—compliance with the World Health Organization’s physical activity guidelines. Preventive medicine. 2015;81:333–8.

30. H Migueles JH, Cadenas-Sánchez C, Alcantara JMA, Leal-Martín J, Mañas A, Ara I, et al. Calibration and cross-validation of accelerometer cut-points to classify sedentary time and physical activity from hip and non-dominant and dominant wrists in older adults. Sensors. 2021;21(10).

31. Moreno-Llamas A, Garcia-Mayor J, De la Cruz-Sanchez E. Concurrent and Convergent Validity of a Single, Brief Question for Physical Activity Assessment. Int J Environ Res Public Health. 2020;17(6):18.

32. Sanders GJ, Boddy LM, Sparks SA, Curry WB, Roe B, Kaehne A, et al. Evaluation of wrist and hip sedentary behaviour and moderate-to-vigorous physical activity raw acceleration cutpoints in older adults. Journal of Sports Sciences. 2019;37(11):1270–9.

33. Zwolinsky S, McKenna J, Pringle A, Widdop P, Griffiths C. Agreement Between The Single-item Measure And IPAQ Using Contemporary Physical Activity Recommendations. Medicine and Science in Sports and Exercise. 2015;47(5):112-.

34. Fynn JF, Hardeman W, Milton K, Jones A. Exploring influences on evaluation practice: a case study of a national physical activity programme. International Journal of Behavioral Nutrition and Physical Activity. 2021;18(1):31.

35. Kuronuma K, Rozanski A, Han DH, Park R, Tomasino GF, Hayes SW, et al. Use of a single-item exercise questionnaire predicts prognostic risk among patients undergoing stress PET-MPI. Journal of Nuclear Cardiology. 2024;41.

36. Dairo YM, Collett J, Dawes H. Development of a single-item physical activity intention measure for adults with intellectual disabilities: Evidence of validity and reliability. Disability and Health Journal. 2024;17(4).

